# Insight and Suicidality in First Episode Psychosis: The Mediating Role of Depression

**DOI:** 10.1101/2024.07.24.24310927

**Authors:** Sümeyra N. Tayfur, Zhiqian Song, Fangyong Li, Hadar Hazan, Toni Gibbs-Dean, Deepa Purushothaman, Sneha Karmani, Javier Ponce Terashima, Cenk Tek, Vinod H. Srihari

**Affiliations:** Program for Specialized Treatment Early in Psychosis (STEP), Yale University School of Medicine, Department of Psychiatry, New Haven, CT, USA; Yale Center for Analytical Sciences (YCAS), Yale School of Public Health, New Haven, CT, USA; Yale University School of Medicine, Department of Psychiatry, New Haven, CT, USA

## Abstract

Understanding the relationship between insight, depression, and suicidality in first-episode psychosis (FEP) is crucial for improving clinical outcomes and preventing suicide during early treatment stages. This longitudinal cohort study examined 264 participants enrolled in coordinated specialty care (CSC) services for FEP to investigate how insight and depression at admission impact suicidality at 6 and 12 months, assess depression’s mediating role between insight and suicidality, and evaluate the persistence of depression over time. Regression analyses assessed the relationships among these variables, while mediation analyses explored depression’s mediating effect. Significant predictors of suicidality at 6 months included insight (OR 0.71, 95% CI: 0.53 - 0.94), depression (OR 5.40, 95% CI: 2.45 - 12.61), and previous suicide attempts (OR 2.91, 95% CI: 1.21 - 7.00). At 12 months, insight (OR 0.70, 95% CI: 0.52 - 0.92) and depression (OR 2.82, 95% CI: 1.26 - 6.50) remained significant. Depression mediated 27.32% of the effect of insight on suicidality at 6 months and 19.76% at 12 months. Despite a general decrease in depressive symptoms, a subset of participants remained persistently depressed. The study highlights the significant mediating role of depression in the relationship between insight and suicidality, with depression emerging as the strongest predictor of suicidality. Early detection and treatment of depression in FEP should be prioritized, and further research should focus on targeted interventions within CSC.

## 1. Introduction

Suicide remains a significant concern for individuals experiencing first-episode psychosis (FEP), particularly within the first year, where the risk is elevated by 60% compared to later stages of the illness (Bornheimer, 2019). It is more common among females than males (Austad et al., 2015). Risk factors highlighted for suicidality in FEP include longer duration of untreated psychosis (DUP), positive symptoms of psychosis, previous suicide attempts, substance use, and lower functioning at entry (Bornheimer, 2019; Coentre et al., 2017). Impaired awareness of symptoms and/or disorder, commonly observed in schizophrenia, is associated with a range of both negative and positive outcomes, a phenomenon known as the “insight paradox” (Davis et al., 2020). While better insight is linked to better treatment adherence and improved social functioning, it is also associated with increased suicidality, depression, and a lower quality of life (Davis et al., 2020; Barrett et al., 2010; Bornheimer et al., 2021). Being female has also been linked to better insight (McEvoy et al., 2006). Approximately 47% of FEP patients experience suicidal ideation, and those who attempt suicide tend to have better insight (Barrett et al., 2010; Foley et al., 2008). However, some studies report no relationship (Restifo et al., 2009) or even a protective effect (Bourgeois et al., 2004). These mixed findings, and the ongoing debate about whether insight is a state or trait further complicate efforts to understand, measure, and modify insight in FEP (e.g., Parellada et al., 2009).

Depression is consistently associated with increased long-term risk for suicide in FEP, surpassing even the impact of command hallucinations (Coentre et al., 2017; McGinty et al., 2018). It is also linked to other adverse outcomes such as worse functioning, higher relapse risk, and lower remission rates (McGinty et al., 2018; McGinty and Upthegrove, 2020). Depression is particularly prevalent at the onset of illness, with rates around 50%, yet it remains underrecognized and undertreated (Bashir et al., 2022; Sönmez et al., 2013; Sönmez et al., 2016). There is a prevailing belief that better insight might lead to depression, thereby increasing suicide risk. However, some argue that insight is crucial for managing the illness and could potentially lower suicide risk by improving treatment adherence. Given the importance of suicide risk in FEP, we’ve explored a) whether insight and depression at entry into a coordinated specialty care (CSC) service predict suicidality at 6 and 12-months, b) whether depression mediates any relationship between baseline insight and suicidality at 6 and 12-months; and c) whether depression persists over 6 and 12-months.

## 2. Methods

### 2.1. Population and study design

Data were drawn from two equivalent CSC services (Feb 1, 2014 - Jan 31, 2019) located in Boston, Massachusetts (Prevention and Recovery in Early Psychosis, PREP) and New Haven, Connecticut (Clinic for Specialized Treatment Early in Psychosis, STEP). Assessments were collected as part of a controlled study of early detection, and all study procedures were approved and monitored by the Institutional Review Boards of Yale University, Beth Israel Deaconess Medical Center, and the Massachusetts Department of Mental Health. Inclusion criteria comprised individuals with a diagnosis of first episode of primary non-affective psychosis, aged between 16 and 35 years and living in the clinic catchment area. Exclusion criteria included psychosis secondary to a previously established medical illness, affective or substance use disorder, and those unable to communicate in English or provide meaningful informed consent (due to cognitive limitations or pre-trial mandates for treatment), and a DUP exceeding 3 years. All participants provided informed consent. Data were collected by trained raters at three time points post-enrollment to the CSC: baseline, 6 months, and 12 months. STEP and PREP recruited comparable, diverse samples reflective of local demography (Srihari et al., 2022).

### 2.2. Measures

Insight was measured with the Positive and Negative Syndrome Scale (PANSS) (Kay et al., 1987). The item G12 from PANSS is frequently used to measure insight, and includes assessment of awareness of mental disorder, its social consequences, and the need for treatment (Barrett et al., 2010; Amore et al., 2020). The G12 severity rating ranges from 1 (no impairment) to 7 (extreme impairment). A higher score indicates poor insight.

Depression was measured using the Calgary Depression Scale for Schizophrenia (CDSS), used extensively to assess the severity of depression in schizophrenia (Addington et al., 1992). Depression is measured in the past week using eight of the nine items of the CDSS, including depression, hopelessness, self-depreciation, guilty ideas of reference, pathological guilt, morning depression, early wakening, and observed depression. The suicide item was removed from the score in this analysis. Items are graded on a 4-point Likert type scale with higher scores indicating more severe symptoms. A cut-off of ≥6 was used to identify those with depression (Sönmez et al., 2016; Chang et al., 2015; Phahladira et al., 2021).

Suicidality was assessed using the Columbia Suicide Severity Rating Scale (C-SSRS) which is designed to measure suicidal ideation and behaviors through two subsets of items (Posner et al., 2011). The first subset focuses on suicidal ideation, with questions such as 1) “*Have you wished you were dead or wished you could go to sleep and not wake up?*” and 2) “*Have you actually had any thoughts of killing yourself?*” If respondents answer “yes” to either of these questions, more detailed queries are made to identify the presence of suicidal behavior (i.e., actual and aborted suicide attempts). We defined suicidality as the presence of either suicidal ideation or behavior and categorized participants into two groups: those with suicidal ideation and/or behavior, and those without either. Suicidal ideation is consistently recognized as a precursor to suicidal behavior (attempt and completion), justifying its inclusion (Bornheimer, 2019).

Positive symptoms were measured with the PANSS using the sum of items P1 (Delusions) to P7 (Hostility) with a range from 7 to 49. Substance use was measured with items from the Alcohol Use Scale (AUS) and Drug Use Scale (DUS) (Drake et al., 1996). Tobacco, alcohol, and cannabis use were rated from 1 (Abstinent) to 5 (Dependence with Institutionalization) and subjects were grouped into three categories as “abstinent”, “use without impairment” and “use disorder”. Functioning was measured with the Global Assessment of Functioning (GAF) scale (Hall, 1995). GAF measures overall levels of current and recent (12 months prior) psychological, social, and occupational functioning and impairment. The score ranges from 1 (severe impairment) to 100 (high functioning). The current score was used in the analysis. Previous suicide attempts at admission were measured using the suicidal behavior section of the C-SSRS and subjects were categorized as positive if they reported actual or aborted suicide attempts. DUP was defined as the time from onset of psychosis to entry into CSC service. The Structured Interview for Psychosis-risk Syndromes (SIPS) was used to establish the transition date to psychosis (McGlashan et al., 2001). The onset of psychosis was defined as meeting the Presence of Psychotic Syndrome (POPS) criteria in SIPS (Srihari et al., 2022).

### 2.3. Statistical analysis

Patient demographics and clinical characteristics were summarized using descriptive statistics. Logistic and linear regression models were used to assess the associations between variables. Mediation analysis was conducted to estimate the role of depression in the association between insight and suicidality using the method by Baron and Kenny (Baron and Kenny, 1986). First, we examined if baseline insight predicted 6-month and 12-month suicidality. Second, we examined if baseline insight correlated with baseline depression. Third, we ran multivariable regression including depression in the model where baseline insight was a predictor. Lastly, the Sobel test was used to estimate what percentage of the total effect of insight on suicidality was mediated by depression. Additional exploratory analyses were conducted to evaluate for effect modification by demographic variables. Analyses were performed using SAS software version 9.4 (Cary, NC). Statistical significance was set at two-sided p<0.05. Figure 1 illustrates the hypothesized model.

**Figure 1.**
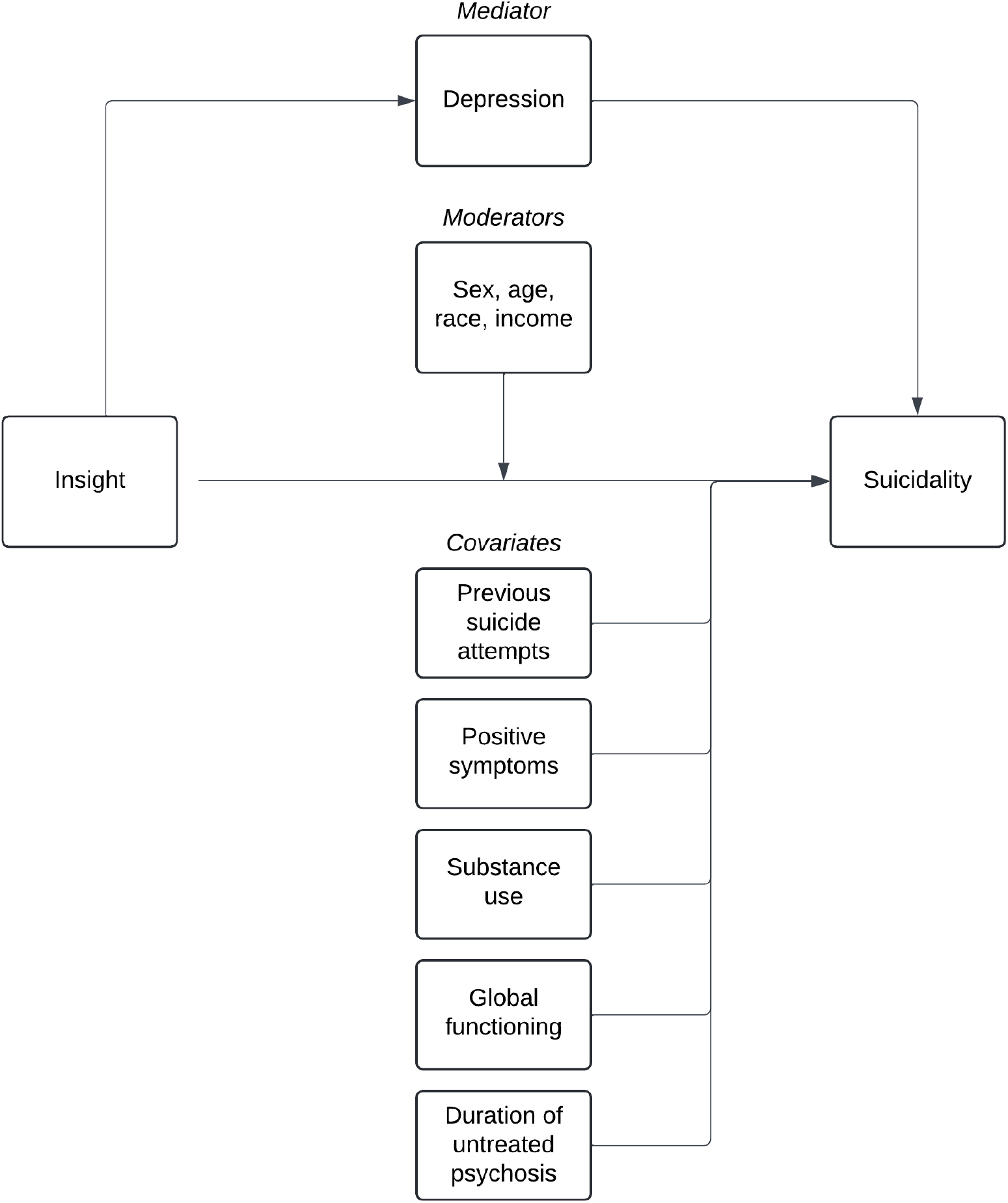
Hypothesized model for the relationship between insight, depression, and suicidality.

## 3. Results

### 3.1. Patient characteristics

A total of 264 participants (188 male and 76 female) were included in the analysis (*M*_*age*_= 22y, *SD* =3.6) (Table 1). They were predominantly Black (44.7%), and the majority (71.6%) had a personal income of less than $10,000. The mean positive symptoms score at admission was 19 (5.9) and 32.8 (11.2) for functioning. At admission, 20.8% of the sample (n=55) reported previous suicide attempts. The majority were not smoking (62.1%) or drinking (47%) or were drinking without impairment 119 (45.1%). Although the majority were not using cannabis (49%), a significant proportion had cannabis use disorder (33.7%). Median DUP was 224 days (IQR: 67-531) (Table 1). The median for insight at admission was 3 (1-4), and 2 (1-4) for 6-months, and 3 (1-4) for 12-months. More than half of the sample met criteria for suicidality at admission (146; 55.3%). This reduced to 19% (n=50) at 6-months and 17.4% (n=46) at 12-months. More than a third of the sample met criteria for depression at admission (101; 38%). This reduced to 18.6% (n=49) at 6-months and 18.2% (n=48) at 12-months. Depression was recurrent in 28 individuals (10.6%) who were also depressed at 6 or 12 months, and persistent in 18 individuals (6.8%) who were depressed at both 6 and 12 months accounting for 17.82% of those who were depressed at baseline.

**Table 1.** Sample demographic and clinical characteristics.

|  | N (%) |
| --- | --- |
| **Sex** |  |
| Male | 188 (71.2%) |
| Female | 76 (28.8%) |
| **Age (mean, SD)** | 22.2 (3.6) |
| **Racial background** |  |
| White (European) | 83 (31.4%) |
| Black | 118 (44.7%) |
| Interracial | 39 (14.8%) |
| Other | 24 (9.1%) |
| **Personal income** |  |
| Less than \$10,000 | 189 (71.6%) |
| More than \$10,000 | 58 (22.0%) |
| **Baseline positive symptoms (mean, SD)** | 19 (5.9) |
| **Previous suicide attempts (yes)** | 55 (20.8%) |
| **Baseline substance use** |  |
| Tobacco (abstinent/use/disorder) | 164 (62.1%) / 97 (36.7%) / 3 (1.1%) |
| Alcohol (abstinent/use/disorder) | 124 (47%) / 119 (45.1%) / 21 (8%) |
| Cannabis (abstinent/use/disorder) | 129 (49%) / 46 (17.4%) / 89 (33.7%) |
| **Baseline functioning (mean, SD)** | 32.8 (11.2) |
| **DUP in days (median, IQR)** | 224 (67-531) |
| **Baseline insight (median, IQR)** | 3 (1-4) |
| **6-month insight (median, IQR)** | 2 (1-4) |
| **12-month insight (median, IQR)** | 3 (1-4) |
| **Baseline depression (yes)** | 101 (38.3%) |
| **6-month depression (yes)** | 49 (18.6%) |
| **12-month depression (yes)** | 48 (18.2%) |
| **Baseline suicidality (yes)** | 146 (55.3%) |
| **6-month suicidality (yes)** | 50 (19%) |
| **12-month suicidality (yes)** | 46 (17.4%) |

**Table 2.** Association of baseline variables with suicidality at follow-up.

| Parameter | Unadjusted |  | Adjusted |  |
| --- | --- | --- | --- | --- |
|  | OR (95% CI) | p value | OR (95% CI) | p value |
| **Suicide at 6-months** |  |  |  |  |
| **Insight** | 0.63 (0.50, 0.79) | <.001 | 0.71 (0.53, 0.94) | 0.017 |
| **Depression** | 7.51 (3.63, 15.5) | <.001 | 5.40 (2.45, 12.61) | 0.001 |
| **Previous suicide attempts** | 3.65 (1.79, 7.44) | <.001 | 2.91 (1.21, 7.00) | 0.016 |
| **Positive symptoms** | 1.01 (0.95, 1.06) | 0.826 | 1.00 (0.92, 1.08) | 0.908 |
| **Smoking (abuse/use vs abstinent)** | 0.67 (0.34, 1.32) | 0.243 | 0.56 (0.22, 1.37) | 0.215 |
| **Alcohol (abuse/use vs abstinent)** | 0.94 (0.50, 1.77) | 0.856 | 1.09 (0.42, 2.85) | 0.864 |
| **Cannabis (use vs abstinent)** | 0.70 (0.28, 1.77) | 0.364 | 0.82 (0.23, 2.70) | 0.460 |
| **Cannabis (abuse vs abstinent)** | 1.11 (0.56, 2.23) | 0.364 | 1.54 (0.56, 4.27) | 0.258 |
| **Global functioning** | 0.98 (0.95, 1.01) | 0.203 | 0.99 (0.95, 1.03) | 0.762 |
| **DUP (per month)** | 1.00 (1.00, 1.00) | 0.886 | 1.01 (0.97, 1.05) | 0.653 |
| **Suicide at 12-months** |  |  |  |  |
| **Insight** | 0.62 (0.49, 0.79) | <.001 | 0.70 (0.52, 0.92) | 0.012 |
| **Depression** | 4.09 (2.04, 8.23) | <.001 | 2.82 (1.26, 6.50) | 0.013 |
| **Previous suicide attempts** | 3.40 (1.62, 7.11) | 0.001 | 2.16 (0.88, 5.19) | 0.088 |
| **Positive symptoms** | 1.02 (0.96, 1.07) | 0.585 | 0.99 (0.91, 1.07) | 0.741 |
| **Smoking (abuse/use vs abstinent)** | 1.33 (0.69, 2.58) | 0.393 | 1.37 (0.58, 3.24) | 0.475 |
| **Alcohol (abuse/use vs abstinent)** | 0.97 (0.50, 1.86) | 0.917 | 0.81 (0.33, 2.00) | 0.638 |
| **Cannabis (use vs abstinent)** | 0.93 (0.36, 2.42) | 0.819 | 1.36 (0.39, 4.46) | 0.887 |
| **Cannabis (abuse vs abstinent)** | 1.07 (0.52, 2.20) | 0.819 | 1.58 (0.59, 4.33) | 0.511 |
| **Global functioning** | 0.97 (0.94, 1.00) | 0.053 | 0.96 (0.92, 1.00) | 0.082 |
| **DUP (per month)** | 1.00 (1.00, 1.00) | 0.139 | 1.03 (0.99, 1.07) | 0.110 |

### 3.2. Baseline insight predicts 6 and 12-month suicidality

According to univariate regression results, those with worse insight were 30% less likely to report suicidality at 6-months (unadjusted OR 0.63, 95% CI: 0.50 - 0.79) and at 12-months (unadjusted OR 0.62, 95% CI: 0.49 - 0.78). For patients with depression, the risk of suicidality at 6-months was almost 8 times higher compared to those without depression (unadjusted OR 7.51, 95% CI: 3.73 – 16.11), and 4 times higher at 12-months (unadjusted OR 4.10, 95% CI: 2.07 – 8.42). After controlling for covariates, the significant predictors of suicidality at 6 months were insight (OR 0.71, 95% CI: 0.53 - 0.94), depression (OR 5.40, 95% CI: 2.45 - 12.61) and previous suicide attempts (OR 2.91, 95% CI: 1.21 - 7.00). At 12 months, insight (OR 0.70, 95% CI: 0.52 - 0.92) and depression (OR 2.82, 95% CI: 1.26 - 6.50) remained as the only significant predictors showing that depression at admission increased the likelihood of suicidality by threefold. Further adjustment for sample demographics did not significantly change these results.

### 3.3. Association between baseline insight and depression

Baseline insight was significantly, and inversely correlated with baseline depression (r= -0.226, *p*<0.001). Those with worse insight were 30% less likely to be depressed (OR 0.73, 95% CI: 0.61 - 0.86).

### 3.4. Mediation analysis

Among subjects with depression at baseline, 75 (74.3%) had suicidality at baseline, 38 (37.6%) had suicidality at 6-months, 31 (30.7%) had suicidality at 12-months. Among subjects without depression at baseline, 70 (43.8%) had suicidality at baseline, 12 (7.5%) had suicidality at 6-months, 15 (9.4%) had suicidality at 12 months. The mediation analysis showed that depression at admission accounted for 27.32% of the effect of insight on suicidality at 6 months (*p*=0.004) and 19.76% of suicide at 12 months (*p*=0.013). We also checked whether insight mediates the relationship between depression and suicidality at follow-up, showing that the reverse hypothesis is not supported. Baseline insight accounted only for 1.56% of the effect of depression on suicidality at 6-months and 2.36% of suicidality at 12-months.

### 3.5. Moderation analysis

Sex, age, race, and income did not moderate the relationship between baseline insight and suicidality at follow-up. We also explored whether the mediation effect differed by sex. For males, baseline depression accounted for 35.96% of the effect of insight on suicidality at 6 months (*p*=0.007), and 18.07% of suicide at 12 months (*p*=0.045). The results for females were not statistically significant (15.50% at 6-months, *p*=0.338; 20.31% at 12-months, *p*=0.341).

## 4. Discussion

The main findings of the current study indicate that better insight upon entry into CSC services was significantly linked to higher suicidality at 6 and 12 months. However, this association was significantly mediated by depression. Individuals who were depressed at admission also had better insight and were significantly more likely to exhibit suicidality at follow-up. Depression at admission was the strongest predictor of suicidality at one year, with a threefold higher likelihood. While the majority of patients experienced a reduction in depression over time, 7% of individuals were persistently depressed at 6 and 12 months accounting for 18% of those who were depressed at baseline.

Our findings support the insight paradox or the demoralization hypothesis (wherein better insight is associated with higher suicidality) but suggests that this relationship may be substantially mediated by depression. In this sample, the mediating effect of depression was stronger for males vs females which was surprising since females are more likely to have better insight and higher suicidality (Austad et al., 2015; McEvoy et al., 2006). However, the smaller sample size of females delivered an underpowered analysis. Our findings align with previous literature that identifies insight as a risk factor for later suicidality (Bornheimer et al., 2021). Similarly, our findings support the recognition of depression as a significant risk factor for suicidality in FEP (McGinty et al., 2018). Other factors like positive symptoms, substance use, functioning and DUP were not significant which was contradictory to previous studies (Bornheimer, 2019; Coentre et al., 2017). These factors may have been better at admission in this population compared to most FEP individuals who present to usual care. Depression has also been associated with better insight in the literature which our study also supports (Coentre et al., 2017; McEvoy et al., 2006; Amore et al., 2020; Phahladira et al., 2021).

Given that depression strongly predicts suicidality at one year and partially mediates the relationship between insight and suicidality, early intervention services for FEP should prioritize identifying and addressing depression. Although not specifically focused on FEP populations, a review of individuals with schizophrenia supports this finding (López-Moríñigo et al., 2012). Some evidence for the demoralization hypothesis in schizophrenia exists (Bourgeois et al., 2004; Amore et al., 2020). Barrett et al. (2010) found evidence for the insight paradox in FEP but did not confirm full mediation by depression. However, they did not check for partial mediation, and their study was cross-sectional. Another cross-sectional study found depression and previous suicidal history to predict suicidality, mediating any relationship between insight dimensions and suicidality (Massons et al., 2017). Similarly, Bornheimer et al. (2021) found that better insight and higher depression increased the odds of suicidal ideation after baseline but did not investigate a mediation effect.

While over half of the sample experienced suicidality upon admission, 38% exhibited depression. This prevalence aligns with previous studies using the CDSS, reporting depression rates around 40% (Sönmez et al., 2013; Sönmez et al., 2016). Despite high prevalence rates, depression is frequently under-identified by clinicians during FEP, with only 17.7% acknowledging its frequent occurrence. In contrast, up to 54% of clinicians report its presence amongst individuals with chronic psychotic disorders, indicating ongoing challenges in recognizing depression in FEP (Bashir et al., 2022). Current approaches to identifying and managing depression in FEP need improvement (Bashir et al., 2022). Clinicians often struggle to differentiate depression from negative symptoms of psychosis and lack a structured method for identifying depression (Bashir et al., 2022). Despite being the gold standard for assessing depression in schizophrenia, the CDSS is underutilized in clinical settings compared to less specialized scales (Bashir et al., 2022). Incorporating the CDSS into routine practice in early intervention services could improve depression monitoring in FEP, as it demonstrates robust psychometric properties even in individuals at ultra-high risk for psychosis (Rekhi et al., 2018).

In the current study, 11% of the sample were depressed at admission and remained so at either 6 or 12 months, with 7% showing persistent depression at both time points, accounting for 18% of those initially depressed. In patients with FEP, persistent depression ranges between 14-26% at one-year follow-up (Sönmez et al., 2013; Cotton et al., 2012). The relatively smaller percentage found in our study may be attributed to the CSC model of care. The persistence of depression in some individuals underscores the need for continuous monitoring to ensure effective management and improve long-term outcomes. Early improvement in depression predicts better clinical outcomes, particularly psychosis remission (Fraguas et al., 2021). Depression at admission, as measured by the CDSS, was linked to a nearly threefold reduction in the likelihood of psychosis remission over a 10-week follow-up, regardless of initial severity of psychotic symptoms and the type of antipsychotic medication (Fraguas et al., 2021). Therefore, individuals with depression require intensive monitoring and prompt, customized interventions to reduce risks, including suicidality, as demonstrated in the current study.

Although clinicians increasingly recognize the need for early intervention, evidence, and guidelines for treating depression in FEP, whether pharmacological or non-pharmacological, are limited (McGinty et al., 2018; Bashir et al., 2022; Bodoano Sánchez et al., 2023; Sabesan and Palaniyappan, 2020), even for schizophrenia (Dondé et al., 2018). Cognitive Behavioral Therapy (CBT) shows promise in improving negative symptoms and social functioning but is underexplored for depression in FEP (Gaynor et al., 2011; Jauhar et al., 2014; Sönmez et al., 2020). Integrating CBT principles and psychoeducation in the first year after FEP has demonstrated benefits, including reduced suicidal ideation (Pelizza et al., 2022a, 2022b). More RCTs are needed to evaluate antidepressants and other treatments (Sönmez et al., 2020; Pelizza et al., 2022a; Palmer et al., 2023). Clozapine is preferred for its suicide risk reduction (Meltzer et al., 2003), while online therapy and n-3 polyunsaturated fatty acids also show potential (Bodoano Sánchez et al., 2023). Cost-effective adjunct therapies like online behavioral interventions warrant further investigation. Ongoing research should prioritize understanding and treating depression in FEP to enhance clinical outcomes.

Several limitations must be acknowledged. Firstly, our sample is drawn from two CSC sites, potentially limiting generalizability to FEP patients who do not receive this comprehensive and developmentally adapted model of care. Secondly, a smaller sample size limited our ability to detect whether depression mediated the relationship between insight and suicidality in young women with FEP. The observational nature of the study precludes definitive causal inferences linking insight to depression, and suicidality. More longitudinal studies with frequent assessment intervals could provide a clearer temporal understanding of these relationships. The single item measure of insight, while widely used, may have reduced the power of this analysis. Moreover, there is a need to consider the potential impact of other variables such as medication adherence and explore the underlying mechanisms that link insight, depression, and suicidality. Finally, it is also necessary to examine the long-term outcomes of FEP patients with varying levels of insight and depression, to better inform treatment strategies and improve patient prognosis.

## 5. Conclusions

Our study reveals that better insight at CSC admission predicts higher suicidality at 6 and 12 months, and is significantly mediated by depression, which also emerged as the strongest predictor of suicidality. This underscores depression as a critical suicide risk factor in FEP, highlighting the potential for proactive targeting of depression to mitigate this risk. Persistent depression in some FEP individuals underscores the need for continuous monitoring. Addressing depression is crucial not only for reducing suicide risk but also for alleviating suffering and disability. Future research should focus on effective interventions that manage depressive symptoms to improve long-term outcomes in FEP.

## Data Availability

All data produced in the present study are available upon reasonable request to the authors

## References

Addington, D., Addington, J., MaTcka-Tyndale, E., Joyce, J., 1992. Reliability and validity of a depression rating scale for schizophrenics. Schizophr Res. 6, 201–208. 10.1016/0920-9964(92)90003-n.

Amore, M., Murri, M. B., Calcagno, P., Rocca, P., Rossi, A., Aguglia, E., … & Maj, M., 2020. The association between insight and depressive symptoms in schizophrenia: Undirected and Bayesian network analyses. Eur Psychiatry 63, e46. 10.1192%2Fj.eurpsy.2020.45.

Austad, G., Joa, I., Johannessen, J. O., Larsen, T. K., 2015. Gender differences in suicidal behaviour in patients with first-episode psychosis. Early Interv Psychiatry 9, 300–307. 10.1111/eip.12113.

Baron, R.M., Kenny, D.A., 1986. The moderator–mediator variable distinction in social psychological research: Conceptual, strategic, and statistical considerations. Journal of personality and social psychology 51, 1173. https://psycnet.apa.org/doi/10.1037/0022-3514.51.6.1173.

Barrett, E. A., Sundet, K., Faerden, A., Agartz, I., Bratlien, U., Romm, K. L., Mork, E., Rossberg, J.I., Steen, N.E., Andreassen, O.A., Melle, I., 2010. Suicidality in first episode psychosis is associated with insight and negative beliefs about psychosis. Schizophr Res. 123, 257–262. 10.1016/j.schres.2010.07.018.

Bashir, Z., Griffiths, S. L., Upthegrove, R., 2022. Recognition and management of depression in early psychosis. BJPsych Bulletin 46, 83–89. 10.1192/bjb.2021.15.

Bodoano Sanchez, I., Mata Agudo, A., Guerrero-Jimenez, M., Girela Serrano, B., Alvarez Gil, P., Carrillo de Albornoz Calahorro, C.M., Gutierrez-Rojas, L., 2023. Treatment of post-psychotic depression in first-episode psychosis. A systematic review. Nord J Psychiatry 77, 109–117. 10.1080/08039488.2022.2067225.

Bornheimer, L. A., Wojtalik, J. A., Li, J., Cobia, D., Smith, M. J., 2021. Suicidal ideation in first-episode psychosis: Considerations for depression, positive symptoms, clinical insight, and cognition. Schizophr Res. 228, 298–304. 10.1016/j.schres.2020.12.025.

Bornheimer, L. A., 2019. Suicidal ideation in first-episode psychosis (FEP): Examination of symptoms of depression and psychosis among individuals in an early phase of treatment. Suicide Life Threat Behav. 49, 423–431. 10.1111/sltb.12440.

Bourgeois, M., Swendsen, J., Young, F., Amador, X., Pini, S., Cassano, G. B., Lindenmayer, J.P., Hsu, C., Alphs, L., Meltzer, H.Y., InterSePT Study Group., 2004. Awareness of disorder and suicide risk in the treatment of schizophrenia: results of the international suicide prevention trial. Am J Psychiatry 161, 1494–1496. 10.1176/appi.ajp.161.8.1494.

Chang, W. C., Cheung, R., Hui, C. L. M., Lin, J., Chan, S. K. W., Lee, E. H. M., Chen, E. Y. H., 2015. Rate and risk factors of depressive symptoms in Chinese patients presenting with first-episode non-affective psychosis in Hong Kong. Schizophr Res. 168, 99–105. 10.1016/j.schres.2015.07.040.

Coentre, R., Talina, M. C., Góis, C., Figueira, M. L., 2017. Depressive symptoms and suicidal behavior auer first-episode psychosis: a comprehensive systematic review. Psychiatry Res. 253, 240–248. 10.1016/j.psychres.2017.04.010.

Cotton, S.M., Lambert, M., Schimmelmann, B.G., Mackinnon, A., Gleeson, J.F.M., Berk, M., Hides, L., Chanen, A., McGorry, P.D., Conus, P., 2012. Depressive symptoms in first episode schizophrenia spectrum disorder. Schizophr Res. 134, 20–26. 10.1016/j.schres.2011.08.018.

Davis, B. J., Lysaker, P. H., Salyers, M. P., Minor, K. S., 2020. The insight paradox in schizophrenia: a meta-analysis of the relationship between clinical insight and quality of life. Schizophr Res. 223, 9–17. 10.1016/j.schres.2020.07.017.

Dondé, C., Vignaud, P., Poulet, E., Brunelin, J., Haesebaert, F., 2018. Management of depression in patients with schizophrenia spectrum disorders: a critical review of international guidelines. Acta Psychiatr Scand. 138, 289–299. 10.1111/acps.12939.

Drake, R. E., Mueser, K. T., McHugo, G. J., 1996. Clinician rating scales: alcohol use scale (AUS), drug use scale (DUS), and substance abuse treatment scale (SATS). Outcomes assessment in clinical practice 113, 2.

Foley, S., Jackson, D., McWilliams, S., Renwick, L., Sutton, M., Turner, N., Kinsella, A., O’Callaghan, E., 2008. Suicidality prior to presentation in first-episode psychosis. Early Interv Psychiatry 2, 242–246. 10.1111/j.1751-7893.2008.00084.x.

Fraguas, D., Díaz-Caneja, C.M., Pina-Camacho, L., van Rossum, I.W., Baandrup, L., Sommer, I.E., Glenthøj, B., Kahn, R.S., Leucht, S., Arango, C., 2021. The role of depression in the prediction of a “late” remission in first-episode psychosis: An analysis of the OPTiMiSE study. Schizophr Res. 231, 100–107. 10.1016/j.schres.2021.03.010.

Gaynor, K., Dooley, B., Lawlor, E., Lawoyin, R., O’Callaghan, E., 2011. Group cognitive behavioural therapy as a treatment for negative symptoms in first-episode psychosis. Early Interv Psychiatry 5, 168–173. 10.1111/j.1751-7893.2011.00270.x.

Hall, R.C., 1995. Global assessment of functioning: a modified scale. Psychosomatics 36, 267–275.

Jauhar, S., McKenna, P.J., Radua, J., Fung, E., Salvador, R., Laws, K.R., 2014. Cognitive–behavioural therapy for the symptoms of schizophrenia: systematic review and meta-analysis with examination of potential bias. The British Journal of Psychiatry 204, 20–29. 10.1192/bjp.bp.112.116285.

Kay, S. R., Fiszbein, A., Opler, L. A., 1987. The positive and negative syndrome scale (PANSS) for schizophrenia. Schizophr Bull. 13, 261–276. 10.1093/schbul/13.2.261.

López-Moríñigo, J.D., Ramos-Ríos, R., David, A.S., Duha, R., 2012. Insight in schizophrenia and risk of suicide: a systematic update. Compr Psychiatry 53, 313–322. 10.1016/j.comppsych.2011.05.015.

Massons, C., Lopez-Morinigo, J.D., Pousa, E., Ruiz, A., Ochoa, S., Usall, J., Nieto, L., Cobo, J., David, A.S., Duha, R., 2017. Insight and suicidality in psychosis: A cross-sectional study. Psychiatry Res. 252, 147–153. 10.1016/j.psychres.2017.02.059.

McEvoy, J. P., Johnson, J., Perkins, D., Lieberman, J. A., Hamer, R. M., Keefe, R. S., Tohen, M., Glick, I.D., Sharma, T., 2006. Insight in first-episode psychosis. Psychol Med. 36, 1385–1393. 10.1017/S0033291706007793.

McGinty, J., Haque, M. S., Upthegrove, R., 2018. Depression during first episode psychosis and subsequent suicide risk: a systematic review and meta-analysis of longitudinal studies. Schizophr Res. 195, 58–66. 10.1016/j.schres.2017.09.040.

McGinty, J., Upthegrove, R., 2020. Depressive symptoms during first episode psychosis and functional outcome: A systematic review and meta-analysis. Schizophr Res. 218, 14–27. 10.1016/j.schres.2019.12.011.

McGlashan, T.H., Walsh, B.C., Woods, S.W., Addington, J., Cadenhead, K., Cannon, T., Walker, E., 2001. Structured interview for psychosis-risk syndromes. New Haven, CT: Yale School of Medicine.

Meltzer, H.Y., Alphs, L., Green, A.I., Altamura, A.C., Anand, R., Bertoldi, A., Bourgeois, M., Chouinard, G., Islam, M.Z., Kane, J., Krishnan, R., 2003. Clozapine treatment for suicidality in schizophrenia: international suicide prevention trial (InterSePT). Arch Gen Psychiatry 60, 82–91. 10.1001/archpsyc.60.1.82.

Palmer, E.R., Griffiths, S.L., Watkins, B., Weetman, T., Ottridge, R., Patel, S., Woolley, R., Tearne, S., Au, P., Taylor, E., Sadiq, Z., 2023. Antidepressants for the prevention of depression following first-episode psychosis (ADEPP): study protocol for a multi-centre, double-blind, randomised controlled trial. Trials 24, 646. 10.1186/s13063-023-07499-3.

Parellada, M., Fraguas, D., Bombín, I., Otero, S., Castro-Fornieles, J., Baeza, I., Gonzalez-Pinto, A., Graell, M., Soutullo, C., Paya, B., Arango, C., 2009. Insight correlates in child-and adolescent-onset first episodes of psychosis: results from the CAFEPS study. Psychol Med. 39, 1433–1445. 10.1017/s0033291708004868.

Pelizza, L., Quattrone, E., Leuci, E., Paulillo, G., Azzali, S., Pupo, S., Pellegrini, P., 2022a. Anxious-depressive symptoms auer a first episode of schizophrenia: response to treatment and psychopathological considerations from the 2-year “Parma Early Psychosis” program. Psychiatry Res. 317, 114887. 10.1016/j.psychres.2022.114887.

Pelizza, L., Maestri, D., Leuci, E., Quattrone, E., Azzali, S., Paulillo, G., Pellegrini, P., 2022b. Individual psychotherapy can reduce suicidal ideation in first episode psychosis: Further findings from the 2-year follow-up of the ‘parma early psychosis’ programme. Clin Psychol Psychother. 29, 982–989. 10.1002/cpp.2678.

Phahladira, L., Asmal, L., Lückhoff, H.K., du Plessis, S., Scheffler, F., Kilian, S., Smit, R., Buckle, C., Chiliza, B., Emsley, R., 2021. The course and concomitants of depression in first-episode schizophrenia spectrum disorders: A 24-month longitudinal study. Psychiatry Res. 298, 113767. 10.1016/j.psychres.2021.113767.

Posner, K., Brown, G. K., Stanley, B., Brent, D. A., Yershova, K. V., Oquendo, M. A., Currier, G.W., Melvin, G.A., Greenhill, L., Shen, S., Mann, J. J., 2011. The Columbia–Suicide Severity Rating Scale: initial validity and internal consistency findings from three multisite studies with adolescents and adults. Am J Psychiatry 168, 1266–1277. 10.1176/appi.ajp.2011.10111704.

Rekhi, G., Ng, W.Y., Lee, J., 2018. Clinical utility of the Calgary Depression Scale for Schizophrenia in individuals at ultra-high risk of psychosis. Schizophr Res. 193, 423–427. 10.1016/j.schres.2017.06.056.

Restifo, K., Harkavy-Friedman, J. M., Shrout, P. E., 2009. Suicidal behavior in schizophrenia: a test of the demoralization hypothesis. J Nerv Ment Dis. 197, 147–153. 10.1097/nmd.0b013e318199f452.

Sabesan, P., Palaniyappan, L., 2020. Therapeutic abstention in the treatment of depression in first-episode psychosis. J Psychiatry Neurosci. 45, 441–442. 10.1503/jpn.200059.

Sönmez, N., Romm, K.L., Østefjells, T., Grande, M., Jensen, L.H., Hummelen, B., Tesli, M., Melle, I., Røssberg, J.I., 2020. Cognitive behavior therapy in early psychosis with a focus on depression and low self-esteem: A randomized controlled trial. Compr Psychiatry 97, 152–157. 10.1016/j.comppsych.2019.152157.

Sönmez, N., Røssberg, J. I., Evensen, J., Barder, H. E., Haahr, U., ten Velden Hegelstad, W., & Friis, S., 2016. Depressive symptoms in first-episode psychosis: A 10-year follow-up study. Early Interv Psychiatry 10, 227–233. 10.1111/eip.12163.

Sönmez, N., Romm, K. L., Andreasssen, O. A., Melle, I., Røssberg, J. I., 2013. Depressive symptoms in first episode psychosis: a one-year follow-up study. BMC psychiatry 13, 1–9. 10.1186/1471-244x-13-106.

Srihari, V. H., Ferrara, M., Li, F., Kline, E., Gülöksüz, S., Pollard, J. M., … & Keshavan, M. S., 2022. Reducing the duration of untreated psychosis (DUP) in a US community: a quasi-experimental trial. Schizophr Bull. Open 3, sgab057. 10.1093/schizbullopen/sgab057.

